# Calculated grades, predicted grades, forecasted grades and actual A-level grades: Reliability, correlations and predictive validity in medical school applicants, undergraduates, and postgraduates in a time of COVID-19

**DOI:** 10.1101/2020.06.02.20116830

**Authors:** I C McManus, Katherine Woolf, Dave Harrison, Paul A Tiffin, Lewis W Paton, Kevin Yet Fong Cheung, Daniel T. Smith

## Abstract

Calculated A-level grades will replace actual, attained A-levels and other Key Stage 5 qualifications in 2020 in the UK as a result of the COVID-19 pandemic. This paper assesses the likely consequences for medical schools in particular, beginning with an overview of the research literature on predicted grades, concluding that calculated grades are likely to correlate strongly with the predicted grades that schools currently provide on UCAS applications. A notable absence from the literature is evidence on whether predicted grades are better or worse than actual grades in predicting university outcomes. This paper provides such evidence on the reduced predictive validity of predicted A-level grades in comparison with actual A-level grades.

The present study analyses the extensive data on predicted and actual grades which are available in UKMED (United Kingdom Medical Education Database), a large-scale administrative dataset containing longitudinal data from medical school application, through undergraduate and then postgraduate training. In particular, predicted A-level grades as well as actual A-level grades are available, along with undergraduate outcomes and postgraduate outcomes which can be used to assess predictive validity of measures collected at selection. This study looks at two UKMED datasets. In the first dataset we compare actual and predicted A-level grades in 237,030 A-levels taken by medical school applicants between 2010 and 2018. 48.8% of predicted grades were accurate, grades were over-predicted in 44.7% of cases and under-predicted in 6.5% of cases. Some A-level subjects, General Studies in particular, showed a higher degree of over-estimation. Similar over-prediction was found for Extended Project Qualifications, and for SQA Advanced Highers.

The second dataset considered 22,150 18-year old applicants to medical school in 2010 to 2014, who had both predicted and actual A-level grades. 12,600 students entered medical school and had final year outcomes available. In addition there were postgraduate outcomes for 1,340 doctors. Undergraduate outcomes are predicted significantly better by actual, attained A-level grades than by predicted A-level grades, as is also the case for postgraduate outcomes.

Modelling the effect of selecting only on calculated grades suggests that because of the lesser predictive ability of predicted grades, medical school cohorts for the 2020 entry year are likely to under-attain, with 13% more gaining the equivalent of the current lowest decile of performance, and 16% fewer gaining the equivalent of the current top decile, effects which are then likely to follow through into postgraduate training. The problems of predicted/calculated grades can to some extent, although not entirely, be ameliorated, by taking U(K)CAT, BMAT, and perhaps other measures into account to supplement calculated grades. Medical schools will probably also need to consider whether additional teaching is needed for entrants who are struggling, or might have missed out on important aspects of A-level teaching, with extra support being needed, so that standards are maintained.

“… the … exam hall [is] a level playing field for all abilities, races and genders to get the grades they truly worked hard for and in true anonymity (as the examiners marking don’t know you). [… Now we] are being given grades based on mere predictions.” Yasmin Hussein, letter to *The Guardian*, March 29^th^ 2020 [1].

“[Let’s] be honest, this year group will always be different.” Dave Thomson, blogpost on *FFT Educational Lab* [2]

“One headmistress commented that ‘entrance to university on teachers’ estimates may be fraught with unimagined difficulties’. … If there is in the future considerable emphasis on school assessment, some work of calibration is imperatively called for.” James Petch, December 1964[3].

## Background

UK schools, in response to the COVID-19 pandemic, closed on March 20^th^, 2020, and Key Stage 5 [Level 3] public examinations such as A-levels and SQA assessments were cancelled for summer 2020. On April 3^rd^ *Ofqual* (Office of Qualifications and Examinations Regulation) in England announced that A-level, GCSE and other exams under its purview in England would be replaced by *calculated grades*, at the core of which are teachers’ estimates of the grades that their students would attain. The Scottish Qualification Authority (SQA) and other national bodies also announced similar processes for their examinations. Inevitably the announcement of calculated grades (holistic assessments in Scotland) resulted in confusion and uncertainty in examination candidates, particularly those needing A-levels or SQA Advanced Highers^a^ to meet conditional offers for admission to university in autumn 2020. Universities also faced a major problem, having had A-levels taken away, which are, “the single most important bit of information [used in selection]” [4].

Some of the tensions implicit in calculated grades are well seen in the quotation above by Yasmin Hussein, a GCSE student in Birmingham, with its clear emphasis that a key strength of current examination systems such as GCSEs, A-levels and similar qualifications, is the *anonymity* and *externality* of the assessors, who know nothing of the students whose work they are marking. In contrast the replacement of actual grades attained in the exam hall with what Hussein describes as ‘mere predictions’ raises a host of questions, not the least being the possibility of *bias* when judgements are made by teachers^b^. In Spring 2020, as universities enter into the final phases of the annual academic cycle of student selection, the potential problems of using estimated or predicted grades rather than actual or attained grades, need urgent consideration.

This paper will mainly concentrate on medical school applications. Medical education has a range of useful educational measures, including admissions tests during selection, and outcomes at the end of undergraduate training, which are linked together through UKMED (United Kingdom Medical Education Database; https://www.ukmed.ac.uk/). UKMED provides a sophisticated platform for assessing predictive validity in multiple entry cohorts in undergraduate and postgraduate training [5]. The paper should also be read in parallel with a second study from some members of the present team which assesses attitudes and perceptions to calculated grades and other changes in selection of current medical school applicants in the UKMACS (UK Medical Applicants Cohort Study) [6].

Fundamental questions about selection in 2020 concern the likely nature of calculated grades, and the extent to which they will predict outcomes to the same extent as currently do *actual or attained grades*. The discussion will involve actual grades, predicted grades, calculated grades, centre assessment grades and forecasted A-level grades, which are summarised in a box below.

### Calculated grades

The status of calculated grades needs to be made clear. *Ofqual* states firmly that,

> “The grades awarded to students will have equal status to the grades awarded in other years and should be treated in this way by universities, colleges and employers. On the results slips and certificates, grades will be reported in the same way as in previous years”. [7], p.6.

The decisions of *Ofqual* in this case are in effect governmental decrees, being supported by Ministerial statement, and universities and other bodies have little choice therefore but to abide by them, although that does not mean that other factors may not need to be taken into account in some cases, as often occurs when applicants do not attain the grades in conditional offers.

None of the above means that calculated grades actually *will be* equivalent to conventional attained grades. Calculated grades will not actually *be* attained grades, they may well behave differently to attained grades, and in measurement terms they actually *are not* attained grades, even though in administrative and even in legal terms, by fiat, they have to be treated as equivalent^c^. From the perspective of educational research, although the key issue is the extent to which calculated grades actually will or can behave in an identical way to attained grades.

##### Actual, predicted, calculated, centre assessment, and forecasted A-level grades

**Actual or attained grades**. The grades awarded by examination boards/awarding organisations based on written and other assessments which are set and marked externally. Typically sat in *May and June of year 13*, with results announced in *mid-August*.

**Predicted grades**. Teacher estimates of the likely attained grades of candidates, provided to UCAS in the *first term of year 13*, and by *October 15^th^* for medical and some other applicants.

**Calculated grades**. The final grades to be provided for candidates by exam boards for Summer 2020 assessments, in the absence of attained grades. Based on ‘centre assessment grades’, with final calculated grades involving standardisation by exam boards. Calculated grades, “will have equal status to the grades awarded in other years and should be treated in this way by universities, colleges and employers” (Ofqual).

**Centre assessment grades**. Used in the production of Calculated Grades (see above). To be provided by examination centres (typically, schools) between 1st and 12^th^ of June, 2020, consisting of grade estimates and candidate rankings within examination centres.

**Forecasted grades**. Prior to 2015, teachers, in *May of Year 13*, provided to exam boards a forecast of the likely grades of candidates along with rankings. Forecasted grades therefore take place later in the academic cycle than predicted grades, close to the time examinations are actually sat.

In April 2020 Ofqual issued guidance on how *calculated grades* would be provided for candidates for whom examinations have been cancelled. Essentially, teachers will be required, for individual candidates taking individual subjects within a *candidate assessment centre*, to estimate *grades for* candidates, and then to *rank order* candidates within grades, to produce *centre assessment grades*.

A statistical standardisation process will then be carried out centrally. Ranking is needed because standardisation, “will need more granular information than the grade alone” ([7] p.7), presumably to break ties at grade boundaries which occur because of standardisation. Standardisation, to produce *calculated grades*, will take into account the typical distribution of results from that centre for that subject in the three previous years, along with aggregated centre data on SATS and previous exam attainment as in GCSEs. This approach is consistent with Ofqual’s approach to standard-setting. Following Cresswell [8], Ofqual has argued that during times of change in assessments, and perhaps more generally, there should be a shift away from “comparable performance” (i.e. criterion-referencing), and that there is an “ethical imperative” to use “comparable outcomes” (i.e. norm-referencing) to minimise advantages and disadvantages to the first cohort taking a new assessment, as perhaps also for later cohorts as teachers improve at teaching new assessments [9].

Ofqual says that centre assessment grades, the core of calculated grades, “are not the same as … predicted grades provided to UCAS in support of university applications” [10], (p.7). Predicted grades in particular are provided by schools in October of year 13 and centre assessment grades in May/June of year 13, seven months later, when Ofqual says that teachers should also consider classwork, bookwork, assignments, mock exams and previous examinations such as AS-levels (taken only by a minority of candidates now), but should *not* include GCSE results, or any student work carried out after 20^th^ March. Whether centre assessment grades, or the closely related calculated grades, will be fundamentally different from predicted grades is ultimately an empirical question, which should be answerable when UCAS data for 2020 are available for medical school applicants in UKMED. In the meantime, it seems a reasonable assumption, except for a small proportion of candidates who have improved dramatically from October 2019 to March 2020, that centre assessment grades and hence calculated grades will probably correlate highly with earlier predicted grades. Predicted grades, which have been collected for decades, should therefore act as a reasonable proxy in research terms for calculated grades.

### The rationale for using A-level grades in selection

Stepping back slightly it is worth revisiting the reasons that A-levels exist and why universities use them in selection. A-levels assess at least three things: subject knowledge, intellectual ability, and study habits such as conscientiousness [11]. Knowledge and understanding of, say, chemistry is probably necessary for high level study of medical science and medicine, to which it provides an underpinning, and experience suggests that students without such knowledge may have problems. A-levels also provide evidence for a student’s intellectual ability and capability of extended study at a high level. A-levels are regarded as a ‘gold standard’ qualification because of the rigour and objectivity of their setting and marking. Their measurement is therefore *reliable*, and the presumption is that they are also *valid*, in the many senses of that word [12-14], and as a result are *unbiased*. A crucial assumption is of *predictive validity*, that future outcomes at or after university are higher or better in those who have higher or better A-levels, as found both in predicting degree classes in general [15-17] and medical school performance in particular [18,19]. At the other extreme, A-levels could be compared conceptually with, say, a mere assertion by a friend or colleague that, “Oh yes, they know lots of chemistry”. That is likely neither to be reliable, valid nor unbiased, and hence is a base metal compared with the gold standard of A-levels. The empirical question therefore is where on the continuum from gold to base, lie calculated grades or predicted grades.

The issue of predictive validity has been little discussed in relation to calculated grades, but in a recent *TES (Times Educational Supplement)* survey of teachers, there were comments that, “predictions and staff assessments would never have the same validity as an exam”, so that, “Predictions, past assessment data and mock data is not sufficient, and will never beat the real thing in terms of accuracy.” [20]. The current changes in university selection inevitably mean that difficult policy decisions need to be made by universities and medical schools. Even in the absence of direct, high-quality, evidence, policy-makers still have an obligation to make decisions, and, therefore it is argued, must take theory, related evidence, and so on, into account [21]. This paper will provide both a review of other evidence, and also new results on the related issue of predicted grades, which it will be argued are likely to behave in a way that is similar to calculated grades.

## Review of literature on predicted and forecasted grades

### Predicted grades in university selection

A notable feature of UK universities is that selection mostly takes place before A-levels or equivalent qualifications have been sat, so offers are largely conditional on later attained grades. As a result, UCAS application forms, since their inception in 1964, have included *predicted grades*, estimates by teachers of the A-level grades a student is likely to achieve. Admissions tutors also use other information in making conditional offers. A majority of applicants in England, applying in year 13 for university entry at age 18 will have taken GCSEs at age 16 in year 11, a few still take AS-levels in year 12, some students submit an EPQ (Extended Project Qualification), and UCAS forms also contain candidate statements and school references. Medical school applicants mostly also take admissions tests such as U(K)CAT or BMAT at the beginning of year 13, and many will take part in interviews or MMIs (multiple mini-interviews).

Predicted grades have always been controversial. A House of Commons Briefing Paper in 2019 noted that the UK was unusual among developed countries in using predicted grades^d^, and said that,

> “The use of predicted grades for university admissions has been questioned for a long time. Many critics argue that predicted grades should not be used for university entry because they are not sufficiently accurate and it has been suggested that disadvantaged students in particular lose out under this system.” [22] p.4

Others have suggested that as well as being “biased”, “predicting A-level grades is clearly an imprecise science” [23] (p.418). There have been repeated suggestions over the years, none as yet successful, that predicted grades should be replaced with a PQA (Post-Qualification Applications) system. As Nick Hillman puts it,

> “The oddity of our system is not so much that people apply before receiving their results; the oddity is that huge weight is put on predicted grades, which are notoriously unreliable. … PQA could tackle this…”^e^.

The system of predicted grades is indeed odd, but also odd is the sparsity of academic research into predicted grades, and particularly their effectiveness. The advent of Ofqual’s calculated grades, which are in effect predicted grades carried out in a slightly different way, means there is now an urgent need to know how effective predicted grades are likely to be as a substitute for attained A-level grades. Are they in fact ‘notoriously unreliable’, being ‘mere predictions’, or do they have equivalent predictive validity as attained grades?

### The research literature on predicted grades

As part of the Supplementary Information to this paper we have included a discursive overview of research studies on predicted grades. Here we will merely provide a brief set of comments.

Most studies look at predictions at the level of individual exam subjects, which are graded from E to either A or, from 2010 onwards, A*. The most informative data show all combinations of predicted grades against attained grades (and Figure 1 gives an example for medical school applicants). Many commentators, though, look only at over-predictions (‘optimistic’) and under-predictions (‘pessimistic’). Figure 2 summarises data from five studies of university applicants. Accurate predictions occur in 52% of cases when A is the maximum grade and 17% when A* is the maximum grade (and of course with more categories accuracy is likely to be lower). Grades are mostly over-predicted, in 42% of cases pre-2010 and 73% post-2010, with under-prediction rarer at 7% of cases pre-2010 and 10% post-2010. A number of studies have reported that under-prediction is more common in lower socio-economic groups, non-White applicants, and applicants from state school or further education [24-26]. A statistical issue means such differences are not easy to interpret, as a student predicted A* cannot be under-estimated, and therefore under-estimation will inevitably be more frequent in groups with lower overall levels of attainment.

**Figure 1:**
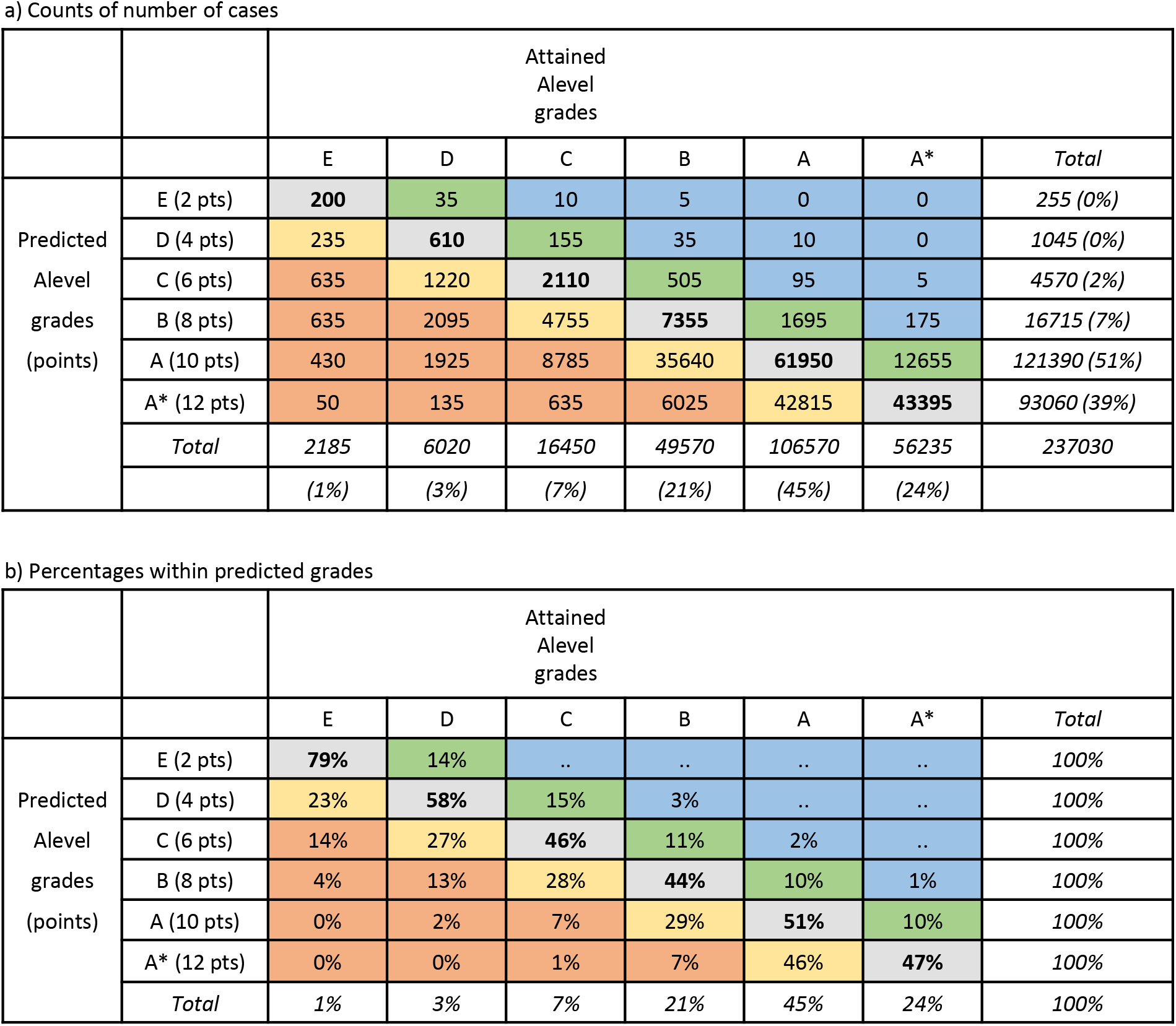
Predicted vs attained A-level grades for individual subjects in applicants to UK medical schools. Accurate predictions are in bold; yellow – over-estimates by 1 grade; orange – over-estimates by 2 grades; green – under-estimates by 1 grade; blue – under-estimates by 2+ grades. a) Counts; b) attained grades as percentages within predicted grades.

**Figure 2:**
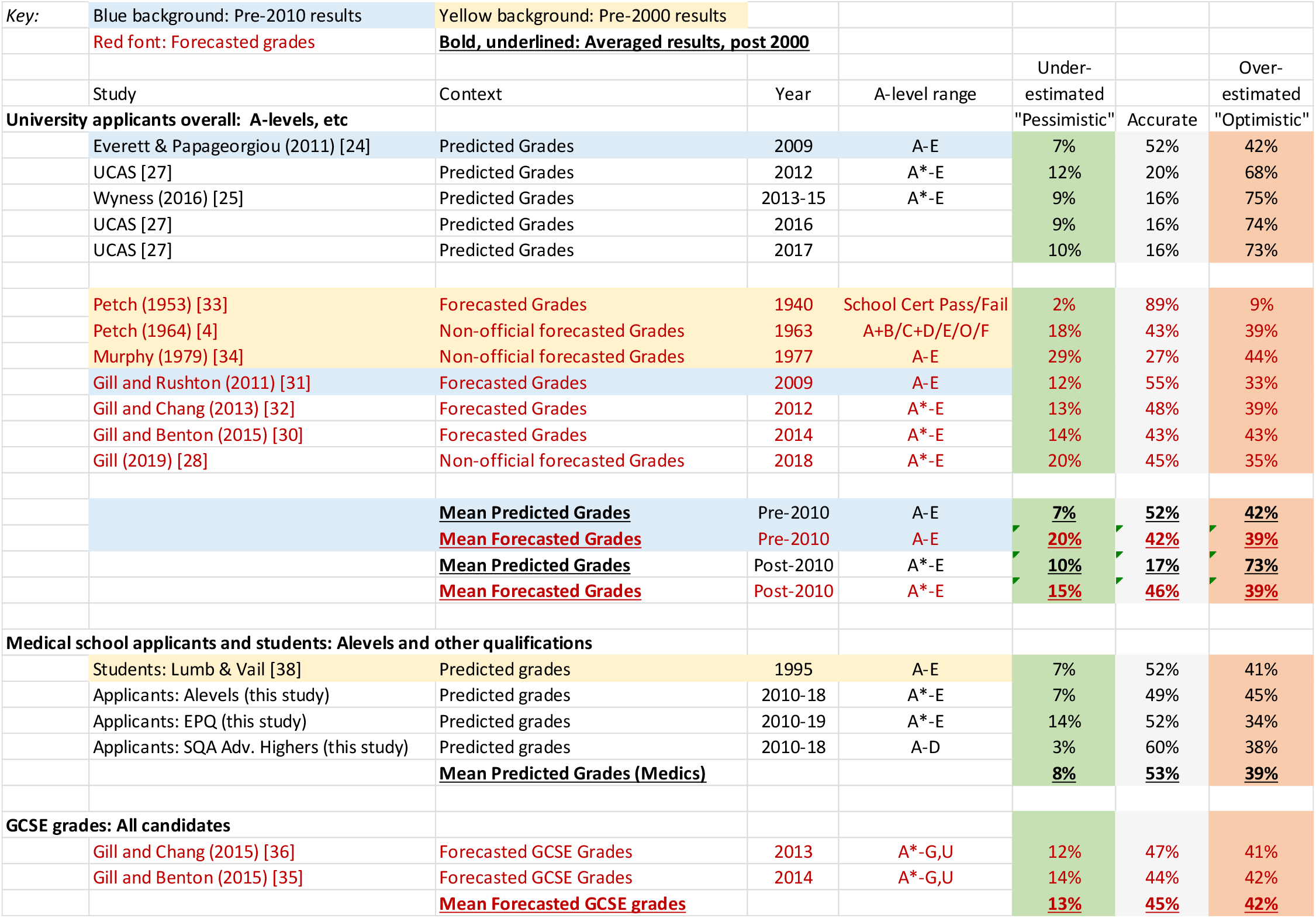
Over-estimates, under-estimate and accurate predicted grades in various studies. Black font: predicted grades; red font: forecasted grades; yellow background: pre-2000; blue background: pre-2010; **<u>bold, underlined</u>**: averaged results post-2000.

Some studies also consider grade-point predictions, the sum of grade scores for the three best attaining subjects, scored A*=12, A=10, B=8, etc^f^.. In particular a large study by UCAS [27] showed that applicants missing their predictions (i.e. they were over-predicted) tended to have lower predicted grades, lower GCSE attainment, were more likely to have taken physics, chemistry, biology and psychology, and were from disadvantaged areas. To some extent the same statistical problems of interpretation apply as with analysis at the level of individual exam subjects. For a number of years UCAS calculated grade-point predictions, and they are included in the P51 data described later.

### What are predicted grades and how are they made?

UCAS says that “A predicted grade is the grade of qualification an applicant’s school or college believes they’re likely to achieve in positive circumstances.”^g^ Later though, the document says predicted grades should be, “**in the best interests of applicants** – fulfilment and success at college or university is the end goal “, and “**aspirational but achievable** – stretching predicted grades are motivational for students, unattainable predicted grades are not” (all emphases in original). Predicted grades should be professional judgements and be data-driven, including the use of, “past Level 2 and Level 3 performance, and/or internal examinations to inform …predictions”.

Few empirical studies have asked how teachers estimate grades, with little major progress since 1964 when Petch said, “Little seems to be known about measures taken by schools to standardize evaluations of pupils” [3] (p.7). Two important exceptions are the studies of Child and Wilson [28] in 2015 and Gill [29] in May 2018, with only the latter published, Gill sending questionnaires to selected OCR Board exam centres concerning Chemistry, English Literature and Psychology exams. Teachers said the most important information used in predicting grades was performance in mock exams, observations of quality of work and commitment, oral presentation, the opinion of other teachers in the same subject, other subjects, and the head of department. Some teachers raised concerns about the lack of high stakes for mock exams which meant that students did not treat them seriously. AS-level grades were an important aid in making predictions. There were also concerns about the loss of AS-levels to help in prediction, as also mentioned elsewhere [30], and that is relevant to 2020 where most candidates will not have taken AS-levels.

### Predicted grades in other Key Stage 5 qualifications than A-levels

Almost all studies on predicted grades have considered A-levels, with a few occasional exceptions looking at GCSEs. We know of no studies on the Extended Project Qualification (EPQ) in England, of Scottish Highers and Advanced Highers, or any other qualifications. The Supplementary Information to this report includes data on both EPQ and SQA examinations.

### Forecasted grades

Until 2015, teachers in the May of Year 13 provided awarding organisations with *forecasted grade*, and those forecasts in part contributed to quality control of grades by the boards. Since forecasted grades are produced five to seven months after predicted grades, and closer to the exam date, they might be expected to be more accurate than predicted grades, being based on better and more recent information. Forecasted grades are important as they are more similar than predicted grades to the proposed calculated grades in the way they are calculated, and it is noted that “they may differ somewhat from the predicted grades sent to UCAS as part of the university application process” [31]. Three formal analyses are available, for candidates in 2009 [32], 2012 [33] and 2014 [31], and four other studies from 1940 [34], 1963 [3], 1977 [35] and 2018 [29] are available, with one post-2000 study before A* grades were introduced and three after (Figure 2). Petch [34] also provides a very early description of forecasted grades, looking at teachers’ predictions of pass or fail in School Certificate examinations in 1940, which also show clear over-prediction.

Forecasted A-level grades are similar in accuracy to predicted grades pre-2010 (42% vs 52%) but are more accurate post-2010 (47% vs 17%), in part due to a large drop in accuracy of later predicted grades. Despite there being *no aspirational or motivational reasons for over-prediction of forecasted grades*, particularly in the 1977 and 2018 studies, over-prediction nevertheless remains as frequent as in predicted grades (pre-2010: 39%; post-2010: 37%) and remains more common than under-prediction (pre-2010: 20%; post-2010 16%). Overall it is possible that calculated grades may be somewhat more accurate than predicted grades, if forecasted grades are taken as a model for calculated grades. Two set of forecasted grades are also available for GCSEs [36,37], and they show similar proportion of over and under-prediction as do results for A-levels. Over-prediction seems to be a feature of all predictions by teachers.

The three non-official studies of forecasted grades also asked teachers to rank-order candidates, a procedure which will also be included in calculated grades. The 1963 data[3] found a median correlation of rankings and exam marks within schools of 0.78, the 1977 data[35] a correlation of 0.66 [35], and the recent 2018 data [29] a correlation of about .82. The three estimates, mean r = 0.75, are somewhat higher than a meta-analytic estimate of .63 (SE =.03) for teachers’ ability to predict academic achievement [38].

The Gill study [29] is also of interest as one teacher commented on the difficulty of providing rankings with 260 students sitting one exam, and the author commented that, “it was easier for smaller centres to make predictions because they know individual students better” (p.42), with it also being the case that responses to the questionnaire were more likely to come from smaller centres. The 1963 study of Petch[3], as well as commenting on “considerable divergencies … in the methods by which estimates were produced” (p.27), as in the variable emphasis put on mock exams, also adds that, “some of the comments from schools suggested that at times there may be a moral ingredient lurking about some of the estimates”(p.28).

The studies of forecasted grades suggest that there is some possibility that calculated grades may be somewhat more accurate than predicted grades, but they also indicate the problems shown by teachers in trying to rank candidates. It also remains possible that examining boards have far more extensive and unpublished data on forecasted grades that they intend to use in assessing the likely effectiveness of calculated grades.

### Applicants to medical school

So far, this paper has been entirely about university applicants across all subjects and the entire range of A-level grades. Only a handful of studies have looked at predicted grades in medical school applicants.

Lumb and Vail pointed out the importance of predicted grades since they determine in large part how shortlisting takes place [39]. In a study of 1995 applicants they found 52% of predictions were accurate, 41% were over-estimated and 7% under-estimated [39], values very similar to those reported in university selection in general (Figure 2).

A study by one of the present team used path modelling to assess the causal inter-relationships of GCSE grades, predicted grades, receipt of an offer, attained A-level grades, and acceptance at medical school [40]. Predicted grades were related to GCSE grades (beta=0.89), and attained A-level grades were predicted by both GCSE grades (beta=0.44) and predicted A-level grades (beta=0.74). The study supports claims that teachers may well be using GCSE grades in part to provide predicted grades, which is perhaps not unreasonable given the clear correlation.

Richardson et al [41] in a very important and seemingly unique study looked at the relative predictive validity of predicted as compared with attained A-level grades. Using a composite outcome of pre-clinical performance they found a minimal correlation with predicted grades (r=.024) compared with a correlation of 0.318 (p<.001) with attained A-level grades. To our knowledge this is the only study of any sort assessing the predictive validity of predicted vs attained A-level grades.

## The present study

Although calculated grades are novel and untested in their details, predicted grades have been around for half a century, and there is also a literature on forecasted grades. This paper will try to answer several empirical questions about predicted grades, for which data are now available in UKMED. Predicted grades will then be used, *faute de mieux*, to make inferences about the likely consequence of using calculated grades.

### Empirical questions to be addressed

- *The relationship between predicted and attained grades in medical school applicants*. Few previous studies have looked in detail at this high-performing group of students. We will also provide brief results on Scottish Highers and Advanced Highers, and the EPQ (Extended Project Qualification), neither of which has been discussed anywhere else to our knowledge.
- *The predictive validity of predicted grades in comparison with attained grades*. A fundamental question concerning calculated grades is whether teacher estimated grades are better or worse at predicting outcomes than are actual A-level grades. The relationship between predicted grades and actual grades cannot answer that question. Instead what matters is how well predicted and actual grades predict subsequent outcomes at the end of undergraduate. The only relatively small study on this of which we are aware in medical students [41] found that only actual grades had predictive validity.

## Method

The method provided here is brief. A fuller description can be found in the Supplementary Information. Overall the project is **UKMEDP112**, approved by the UKMED Research Group in May 2020, with data coming from two separate but related UKMED projects, both of which include predicted grades.

Project **UKMEDP089**, “The UK Medical Applicant Cohort Study: Applications and Outcomes Study”, approved Dec 7^th^, 2018, with Dr Katherine Woolf as principal investigator, is an ongoing analysis of medical student selection as a part of UKMACS (UK Medical Applicant Cohort Study^h^). The data upload of 21^st^ Jan 2020 included detailed information from UCAS and HESA on applicants for medicine from 2007 to 2018.

Project **UKMEDP051**, “A comparison of the properties of BMAT, GAMSAT and UKCAT”, approved Sept 25^th^, 2017, with Dr Paul Tiffin as principal investigator, is an ongoing analysis of the predictive validity of admissions tests and other selection methods such as A-levels and GCSEs in relation to undergraduate and postgraduate attainment. UCAS data are included, although at the time of the present analysis, 16^th^ April 2020, this had not yet included the detailed subject level information available in UKMEDP089^i^. Outcome data for the P51 dataset are extensive, and in particular undergraduate progression data are included, such as UKFPO EPM and SJT, and PSA (Prescribing Safety Assessment).

### Rounding and suppression criteria

Data from HESA are required to be reported using their rounding and suppression criteria (https://www.hesa.ac.uk/about/regulation/data-protection/rounding-and-suppression-anonymise-statistics) and those criteria have been used for all UKMED data. In particular the presence of a zero or a zero percentage may not always mean that there are no individuals in a cell of a table, and all integers are rounded to the nearest 5.

### Ethical approval

Queen Mary Research Ethics Committee, University of London, agreed on 11 November 2015 that there was no need for ethical review of UK Medical Education Database research studies.

### Rounding and suppression criteria

All data from HESA are required to be reported using their rounding and suppression criteria (https://www.hesa.ac.uk/about/regulation/data-protection/rounding-and-suppression-anonymise-statistics) and we have applied those criteria to all UKMED-based tables and values reported here.

## Results

As with the Method section, a fuller description of the results can be found in the Supplementary Information.

**The relationships between predicted and actual grades in medical school applicants**.

### Predicted and actual A-level grades for individual A-level examinations

Figure 1 shows the relationship between predicted and attained A-level grades for 237,030 examinations from 2010 to 2018 (i.e. including A* grades). 39.3% of predicted grades are A* compared with 23.7% of attained grades. Figure 1.a shows predicted grades in relation to attained grades, with bold font for accurate predictions, green and blue shading for under-prediction and orange and red shading for over-prediction. Overall 48.8% of predicted grades are accurate, which is higher than for university applications in general (see Figure 2), reflecting the high proportion of A and A* grades (69%). Over-prediction occurred in 44.7% of cases, and under-prediction in 6.5% of cases. Figure 1.b show the data as percentages. About a half of A* predictions result in an attained A grade, and over a third of predicted A grades result in grade B or lower. Despite over-prediction by about half an A-level grade, predicted and attained grades have a Pearson correlation of r = 0.63.

The question of whether predicted grades are more accurate in private sector schools than public sector, state schools is complex and is discussed in an appendix to the Supplementary Information. There are subtle ceiling effects and non-linearities, but overall the best conclusion seems to be that private sector predictions may seem to be more accurate but that mainly reflects the higher actual grades of students at selective schools (and they have been selected to have higher ability levels). That conclusion is also important as it means that although there is an argument that calculated grades may predict outcomes better than predicted grades because they will have been standardised, predicted grades in effect are already standardised in so far as teachers at selective schools provide higher predicted grades because those candidates in actually do gain higher attained A-level grades. Standardisation by previous school performance is unlikely to change that conclusion.

### Differences between A-level subjects

There is little in the literature on the extent to which different A-level subjects may differ in the accuracy of their predictions, perhaps with different degrees of bias or correlation. Detailed results are presented in the Supplementary Information. Overall, Biology, Chemistry, Maths and Physics are very similar in terms of over-prediction and correlation with actual grades. However General Studies is particularly over-estimated compared with other subjects.

### Extended Project Qualification (EPQ) and SQA Advanced Highers

The Supplementary Information contains information on these qualifications. SQA Advanced Highers, as well as the EPQ, show similar proportions of over-estimation as other qualifications (see Figure 2).

### Reliability of predicted and attained A-level grades

Considering the best three A-level grades, the reliability of an overall score can be calculated from the correlations of the individual subjects. For 66,006 candidates with at least three paired predicted and actual grades, Cronbach’s alpha was 0.827 for actual grades and 0.786 for predicted grades, with a highly significant difference. The difference may in part reflect the higher proportion of A* grades in predicted than actual grades, but may also reflect greater measurement precision in the marking of actual A-levels.

### How reliable are attained A-level grades?

Attained A-level grades, like any behavioural measurement are not perfectly reliable, in the sense that if a candidate took a parallel test containing equivalent but different items it is highly unlikely that they would get exactly the same mark as on the first attempt. They may, for instance, have been lucky (or unlucky) at their first attempt, having questions which they happened to have studied or revised more (or revised less), and so on. Reliability is a technical subject^j^ with many different approaches [42,43]. For continuous measures of raw scores, the reliability can be expressed as a coefficient such as alpha (and in one A-level maths test in 2011, alpha for the full test was about 0.97 [44], although it is suggested that value is unusually high). Boards though do not report raw scores, but instead award grades on a scale such as A* to E. The ‘classification accuracy’ of grades is harder to estimate, and is greater with fewer grade points, wider grade intervals, and a wide spread of candidate ability [44]. There seem to be few published estimates of classification accuracy for A-levels (although they do exist for GCSEs and AS-levels [44]). Estimating classification accuracy for the present high-attaining group of medical school applicants is not easy. A fundamental limit for any applicants is that predicted grades cannot possibly predict actual grades better than attained grades predict themselves (the reliability or classification accuracy). However, from considering the correlation of the three best predicted and actual grades it is unlikely that such a limit has currently been reached. The correlation of actual with predicted grades is .585 (see figure 3), and the alpha reliabilities of .827 for actual grades and .786 for predicted grades (see above). The disattenuated correlation between predicted and actual grades is therefore .585/(V(.827x.786) = 0.726, which is substantially less than one, with predicted grades accounting for only about a half of the true variance present in actual grades. If the disattenuated correlation were close to one then it could be argued that predicted grades were doing as well as they could possibly do given that attained grades are not perfectly reliable, but that is clearly far from the case.

**Figure 3:**
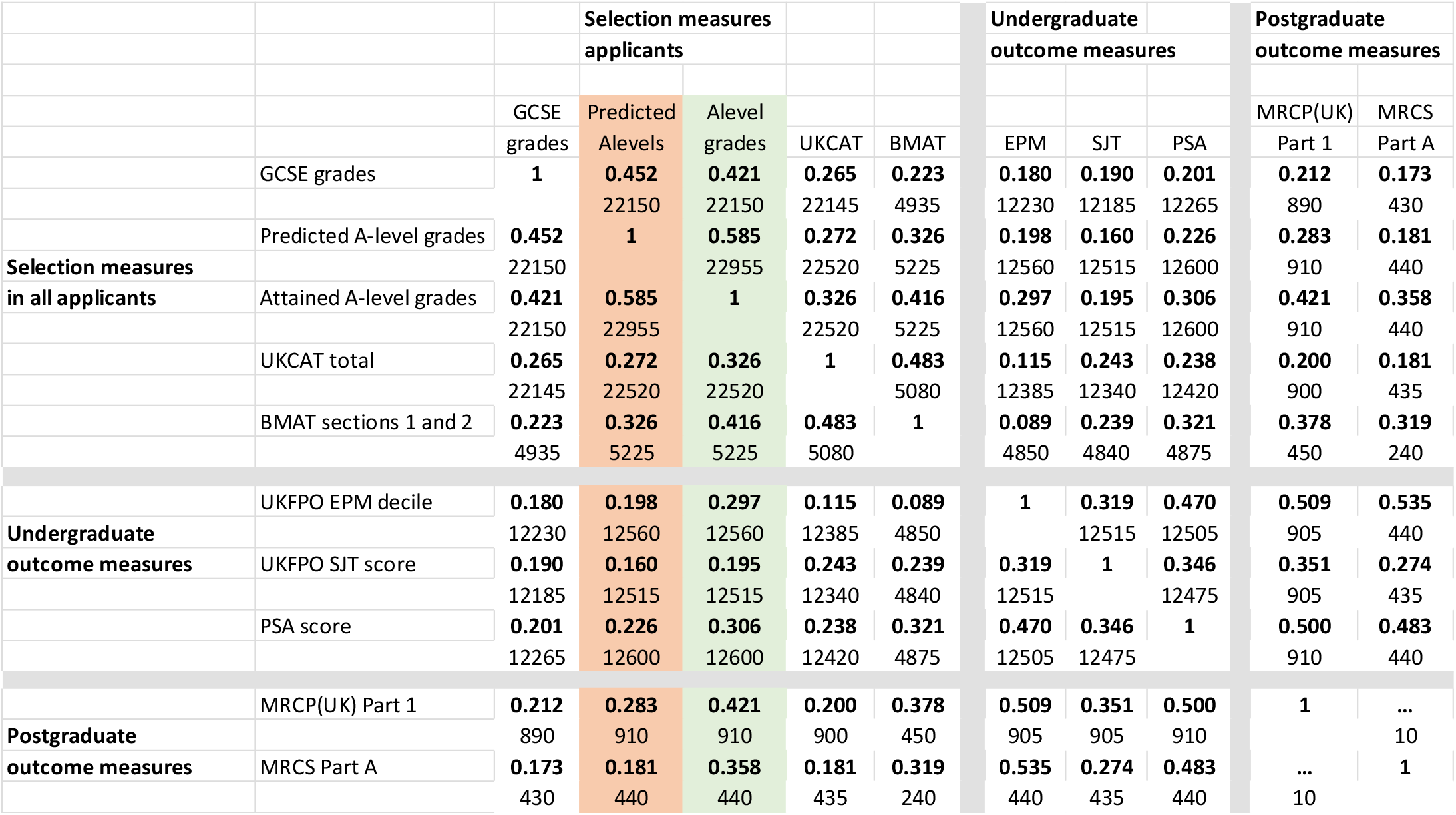
Correlation matrix of selection measures, undergraduate outcome measures, and postgraduate outcome measures (separated by grey lines for clarity). Cells indicate Pearson correlation and N.

### True scores and actual scores

From a theoretical, psychometric, point of view it could be argued that it is neither actual nor predicted grades which need to be estimated for applicants, but their ‘true ability scores’, or the ‘latent scores’, to use the technical expressions, of which predicted and actual grades are but imperfect estimates. In an ideal world that would be the case, and a well-constructed exam tries to get as close as possible to true scores. However, it is not possible to know true scores (and if it were the boards would provide selectors with those scores). Selection itself does not work on true scores but on the actual grades that are written down, by teachers for predicted grades, and as grades on exam result certificates by boards. They are the currency in which transactions are conducted during selection, so that a predicted grade of less than a certain level means a candidate will not get a conditional offer, and likewise too low an actual grade means a candidate holding a conditional offer will be rejected. For that reason it is not strictly the correlation of predicted and actual grades which matters, the two measures being treated as symmetric, but the forward prediction of actual grades from predicted grades, i.e. the actual grades conditional on the predicted grades (as shown in figure 1b).

## Predictive validity of predicted and attained A-level grades in medical students

### Predictive validity in UKMEDP051

The version of the P51 data used here consists entirely of applicants applying to medical schools, but there is also follow-up into undergraduate and postgraduate training. Predicted A-level grades were available only for the UCAS application cycles of 2010 to 2014, and consisted of a single score in the range 4 to 36 points, based on the three highest predictions scored as A*=12, A=10, etc.. The modal score for 38,965 applicants was 30 (equivalent to AAA; mean=31.17; SD= 3.58; Median = 32; 5^th^, 25^th^, 75^th^ and 95^th^ percentiles= 26, 30, 34 and 36). For simplicity the study was restricted to applicants aged 18 in the year of application, who had both predicted and attained A-levels, which also ensured the sample contained only first applications for non-graduate courses, from candidates who had not taken pre-2010 A-levels. Overall, 22,955 applicants were studied. Other selection measures included were GCSEs (mean grade for best eight grades), as well as U(K)CAT and BMAT scores, based on the most recent attempt which for most of these cases was also the first attempt. For simplicity we used the total of the four sub-scores of U(K)CAT, and the total of Section 1 and 2 scores for BMAT.

Follow-up is complicated as application cohorts enter medical school in different years, and spread out in time through medical school. For this UKMED data extract, applicants in 2010 entered the medical register from 2015-18, 2011 applicants in 2016-8, 2012 applicants in 2017-18 and 2013 applicants in 2018. Applicants in 2014 would only qualify in 2019 or after, and the UKMED dataset did not include 2019 data. Some 2010 to 2013 applicants will also qualify after 2018. Again for simplicity, undergraduate outcome measures were restricted to the deciles of the UKFPO’s Educational Performance Measure (EPM), the raw score of the UKFPO’s Situational Judgement Test (SJT), and the score relative to the pass mark of the Prescribing Safety Assessment (PSA), all at first attempt. Relatively few doctors, mostly from the earlier cohorts, had progressed through to postgraduate assessments, but sufficient numbers for analysis were present for MRCP(UK) Part 1 and MRCS Part A, scores being analysed at the first attempt.

EPM, is a complicated measure summarising academic progression through medical school, with individual schools deciding what measures to include [45], and expressed as deciles *within* each school and graduating cohort year. EPM is here used as the main undergraduate outcome measure. Deciles are confusing, as UKFPO scores them in the reverse of the usual order, the first decile being highest performance and the tenth the lowest^k^ (see Figure 5). Here for ease of interpretation we reverse the scoring in what we call *revDecile*, so that higher *revDeciles* indicate higher performance. It should also be remembered that deciles are not an equal interval scale.

Correlations between the measures are summarised in Figure 3. Large differences in Ns reflect some measures being used in applicants during *selection*, and others being outcome measures that are only present in *entrants*, as well as the smaller numbers of doctors who had progressed to postgraduate assessments. The distinction is emphasised by dividing the correlation matrix into three separate parts. Correlations of selection and outcome measures necessarily show *range restriction* because candidates have been selected on the basis of the selection measures, and likewise doctors taking postgraduate examinations may be self-selected for earlier examination performance.

Figure 3 contains much of interest (see Supplementary Information), but the most important question for present purposes is the extent to which Predicted and Attained A-level grades (shown in pink and green in Figure 3) differ in their prediction of the five outcome measures, remembering that undergraduate outcomes are typically five or six years after selection, and postgraduate outcomes are seven or eight years after selection.

Attained A-levels predict EPM with r=0.297 compared with a correlation of only 0.198 for predicted grades. N is large for these correlations and hence the difference, using a test for correlated correlations [46] is highly significant (Z=12.6, p<10^-33^). These raw correlations are inevitably range restricted, and the construct level predictive validity accounting for range restriction and measurement error is likely to be much higher [47]. Multiple regression (see Supplementary Information) suggests that predicted grades may have a small amount of predictive variance which is not shared with attained A-levels. Figure 4 shows mean EPM *revDecile* scores in relation to actual and predicted A-levels. The slope of the line is clearly less for predicted A-levels, showing a less good prediction. It is also clear that attained grades predict well, A*A*A* entrants scoring an average of two deciles higher at the end of the course than those with AAA grades, each extra grade raising average performance by about two-thirds of a decile. In contrast the slope is less for predicted grades, being slightly less than half a decile per predicted A-level grade.

The broad pattern of results is similar for the other undergraduate outcomes, SJT and PSA, and is available in the Supplementary Information.

**Figure 4:**
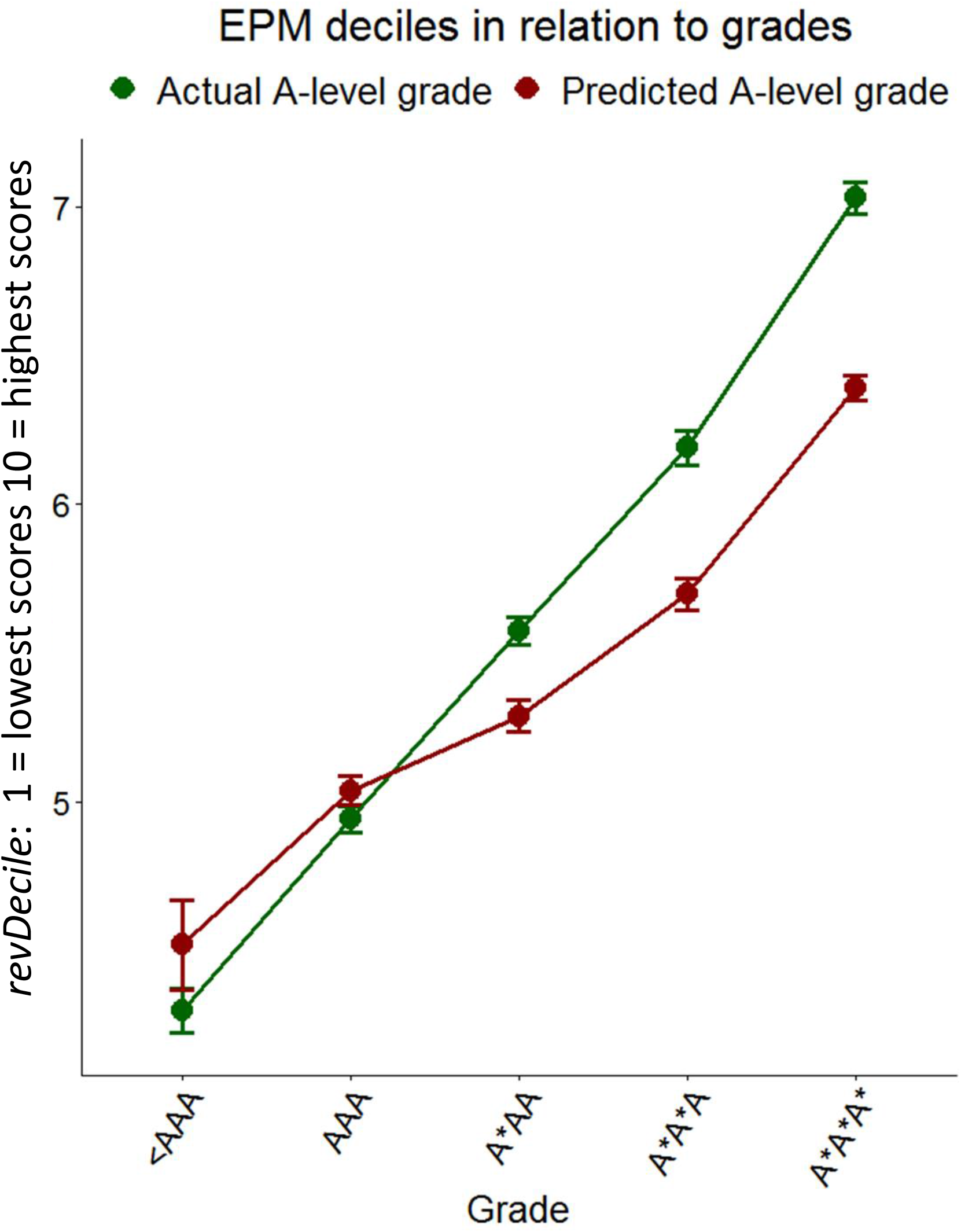
Mean EPM *revDeciles* (95% CI) in relation to actual A-level grades (green) and predicted A-level grades (red)

The two postgraduate outcome measures, MRCP(UK) Part 1 and MRCS Part A, although both based on smaller numbers of doctors, are still significant, actual grades correlating more highly with MRCP(UK) Part 1 (r=.421) than do predicted grades (r=.283; Z= 4.54, p=.000055). Likewise, actual grades correlate more highly with MRCS Part A (r=.421) than do predicted grades (r=.358; Z= 3.67, p=.000238).

There are suggestions that predicted grades may not be equivalent in candidates from state schools and private schools, grades being predicted more accurately in independent schools [24,25]. That is looked at in the Supplementary Information, and while there is clear evidence, as found before in the UKCAT-12 study [48], that private school entrants underperform relative to expectations based on their A-levels, there is no evidence that predicted grades behave differently in candidates from private schools.

A practical question relevant to calculated grades concerns the extent to which, in the absence of attained A-level grades, other selection measures such as GCSEs, U(K)CAT and BMAT can replace the predictive variance of attained A-level grades. That will be considered for EPM where the sample sizes are large. Attained grades alone give R = 0.297, and predicted grades alone give R=.198, accounting for less than half as much outcome variance. Adding GCSEs to a regression model including just predicted grades increases R to .225, and also including U(K)CAT and BMAT increases R to .231, which is though substantially less than the .297 for attained A-levels alone. In the absence of attained A-level grades, prediction is therefore improved by including GCSEs and U(K)CAT or BMAT, although the prediction still falls short of that for actual A-levels alone.

### Modelling the effect of only predicted grades being available for selection

In the context of the 2020 pandemic, an important question is the extent to which future outcomes may change as a result of selection being in terms of calculated grades. Calculated grades themselves cannot be known, but predicted grades are probably a reasonable surrogate for them in the first instance. A modelling exercise was therefore carried out whereby the numbers of students in the various EPM revDeciles were tabulated in relation to predicted grades at five grade levels, 36 pts ≡ A*A*A*, 34 pts ≡ A*A*A, 32 pts ≡ A*AA, 30 pts ≡ AAA and ≤ 28 pts ≡ ≤ AAB, the probability of each decile found for each predicted A-level band. Assuming that selection results in the usual numbers of entrants with grades of A*A*A*, A*A*A, etc, but based on calculated grades rather than actual grades, the expected numbers of students in the various EPM deciles can be found. Figure 5 shows deciles as standard UKFPO deciles (1 = highest), UKFPO scores (43 = highest), and *RevDeciles*, (10 = highest). The blue column shows the actual proportions in the deciles based on attained A-level grades. Note that for various reasons there are not exactly equal proportions in the ten deciles^l^. Based on selection on attained A-level grades there are 7.2% of students in the lowest performing decile, compared with an expected proportion of 8.1% for selection on predicted grades, an increase of 0.9% percentage points, which is a relative increase of 13.0% in the proportion of the lowest decile, with an odds ratio of 1.141 of attaining the lowest decile. For the highest scoring decile, the proportion decreases from 10.1% with actual A-level grades to 8.8% if predicted A-level grades are used, an absolute decrease of 1.4%, and a relative decrease of 13.4% of top deciles, with an odds ratio of 0.853.

**Figure 5:**
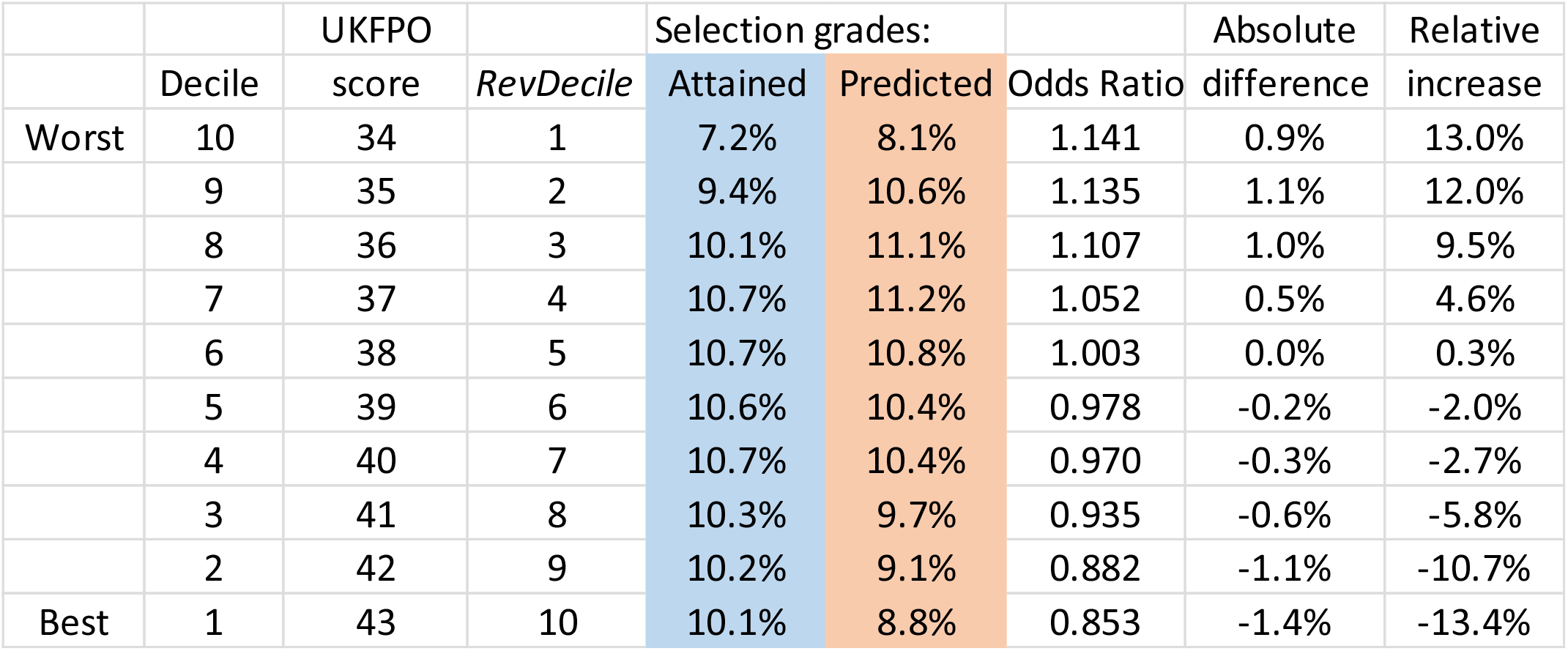
Predicted decile outcomes if selection were on Predicted A-level grades (blue) rather than actual A-level grades (orange).

Of course the above calculations are based on the assumption that the ‘deciles’ for calculated grades are expressed at the same standard as currently. Were the outcomes to be restandardised so that all deciles were equally represented then of course at finals no noticeable difference would be present^m^. However the academic backbone would still be present, and overall poorer performance on statistically equated postgraduate exams would be expected.

## Discussion

Despite predicted grades being a key part of UK university selection at least since the advent in 1964 of UCCA (later UCAS), it is surprising how few answers there have been to key questions about their use. Even simple questions such as whether different A-level subjects are predicted with different accuracy or different degrees of over-estimation are mostly unanswered (in public at least, although awarding bodies or UCAS may know the answers in private), and few of the questions are answered in the context of specific degree courses, such as medicine, where most of the applicants and students are high-flyers. The most important question that seems almost never to have been asked, and certainly not answered, is the fundamental one of whether it is predicted grades or actual grades which are better at predicting outcomes. Petch[3] considers that predicted and actual grades may be fundamentally different, perhaps being “complementary and not contradictory” (p.29), one being about scholarly attitude and the other about examination prowess, primarily because “the school knows the candidate as a pupil, knowledge not available to the examiners”. For Petch, either a zero correlation or a perfect correlation between predicted and actual grades would be problematic, the latter perhaps implying that actual grades might be seen as redundant (p.6). Although scenarios can be envisaged in which the holistic judgments by teachers of predicted grades are better predictors of outcomes, since teachers may know their students better than examiners, the present data make clear that attained grades in fact are far better predictors than predicted grades, which account for only about a third as much outcome variance as attained grades. The immense efforts by exam boards and large numbers of trained markers to refine educational measurements is therefore gratifying and reassuring. Careful measurement does matter. Validation is the bottom line of all educational assessment, and in the present case it is validation against assessment five to eight years down the line from the original A-levels, in both undergraduate and postgraduate assessments. That is strong support for what we have called ‘the academic backbone’, prior attainment providing the underpinning for later attainment, and hence performance at assessments at all stages of training for GCSEs through to medical degrees (and probably on to postgraduate assessments) tend to be correlated [19].

A remaining question is whether there is still some variance in predicted and actual grades which is complementary. A tiny amount, is the answer, predicted grades increasing R by about 0.05 when actual grades are already taken into account. What that variance might be is unclear, and it is possible that it is Petch’s ‘scholarly attitude’. At present though it is worth remembering that *examination* grades at A-level are primarily predicting further *examination* grades at the end of medical school, although EPM scores do include some measures of course work, and practical and clinical skills. If other outcome measures, perhaps to do with communication, caring or other non-cognitive skills were available then predicted grades might show a greater predictive value.

The present data inevitably have some limitations, but they are few. There is little likelihood of bias since complete population samples have been considered, and there is good statistical power with large sample sizes. Inevitably not all outcomes can be considered, mainly because the cohorts analysed have not yet progressed sufficiently through postgraduate training. However those who are included show effects which are highly significant statistically.

Our questions about predicted grades have been asked in the practical context of the cancellation of A-level assessments and their replacement by calculated grades, as a result of the COVID-19 pandemic. It seems reasonable to assume, given the literature on predicted grades, and particularly on forecasted grades, that calculated grades will probably have similar predictive ability to predicted grades, but perhaps be a little more effective due to occurring later in the academic cycle. Such a conclusion would be on firmer ground if exam boards had analysed the predictive validity of the data they had collected on forecasted grades, particularly in comparison with predicted and actual grades. Such data may exist, and if so then they need to be seen. In their absence, the present data may be the best available guesstimates of the likely predictive validity of calculated rather than actual grades.

A further consideration is more general and asks what the broader effects of the COVID-19 pandemic may be on education. Students at all levels of education have had teaching and learning disrupted, often extensively, and that is also true of all stages of medical education. The current cohort of applicants will not be assessed formally at A-level. As well as meaning that they may only have calculated grades, which are likely to be less accurate, they also will have missed out on several other things. Year 13 students may have missed normal schooling from, say, mid-March until mid-May, when they should have taken public examinations, perhaps 30 to 40 school days. Year 12 students, from whom 2021 medical school entrants will be drawn, may miss school from mid-March until mid-July, totalling perhaps 80 school days. Burgess and Sievertsen [49], using data from two studies [50,51], estimate that 60 lost school days results in a reduction in performance of about 6% of a standard deviation, which they say is, “non-trivial”. These effects are likely to differ also by socio-economic background, particularly in effectiveness of home schooling. Applicants not taking A-levels will also suffer from the loss of the enhanced learning that occurs when learners are tested – the ‘testing effect’ – for which a meta-analysis found an effect size of 0.50 [52], which is also non-trivial. Taken overall, 2020 entrants to medical school, and perhaps those in 2021 as well, may perform less well as a result of missing out both on education and on its assessment.

What then are the specific implications of our findings for the use of calculated grades, for medical education and the selection of medical students? Whether our findings will generalise to other university courses is far from clear, not least because very few have the sophisticated undergraduate and postgraduate outcome measures found in medical education, which inevitably will make effects harder to demonstrate, although in principle the effects should still be there.

### For universities and medical schools

Medical schools are skilled in understanding the various data they hold on applicants, and how results ought to look given prior attainment, and they recognise that some sets of results might be skewed, sometimes because of small group sizes. They also recognise that predicted grades often err on the side of generosity. No doubt they will be looking at calculated grades to check whether they are compatible with other data. Nevertheless, calculated grades are largely uncharted territory even for experienced university admissions officers, with many potential uncertainties, particularly in relation to predicting future medical school performance.

A useful thought-experiment is to imagine what would happen were a computer-error accidentally to shuffle all A-level grades, but with the same numbers of A*, A, B grades, etc still being awarded for individual subjects, but without any relation to the grades that candidates should have attained. Administratively, selection would proceed as normal. However instead of medical schools admitting high-achieving entrants with A* and A grades, entrants would have what we might call nominal ‘A*’ and ‘A’ grades, but in reality would have had true grades anywhere from A* to E, with a mode at about B. Since A-levels undoubtedly predict university outcomes, many entrants with lower true grades would therefore probably struggle with course content and exams. For medical schools using criterion-referencing, more students than usual would fail exams. If norm-referencing were used, the usual proportion of students would pass but pass marks would be lower, students would know less at graduation, and the knowledge deficit would then become clear when taking statistically equated postgraduate examinations such as MRCP(UK).

Attained grades within candidates in different A-level subjects normally tend to correlate, so that the reliability of a total points score can be calculated (see above). Grades allocated at random would not however correlate within candidates. Were a medical school to select on total A-level points (say, with a threshold equivalent to AAA) there would still be applicants with that number of points. However because random allocation results in zero correlations of grades within candidates, the variance of total points would be less and candidates with high point scores would be in shorter supply.

Although only a thought-experiment, calculated grades will probably have a partial similarity to randomly allocated grades (and the reliability of predicted grades is indeed less than that of attained grades, because correlations are lower) in that measurement error will have increased. Different subjects will correlate less than usual, and there may be a shortage of qualified candidates, and nominal grades will correlate less with outcomes than usual, regression to the mean making outcomes somewhat poorer than usual. The predictions therefore are qualitatively similar to the pattern of those with random allocation, but less extreme.

A practical question is whether medical schools can do anything to ameliorate possible problems with calculated grades. For those holding conditional offers, a strong argument says there is a contractual agreement between a university and a candidate holding a conditional offer based on certain A-level grades (although a counter-argument might be that the contract implicitly said that payment would be in actual, gold-standard, attained grades, and not in the potentially baser currency of promissory notes based on calculated grades). If indeed there were to be too many candidates meeting their offers, then the precise nature of the contract could become crucial. For candidates not meeting the grades stated in conditional offers, so that there are unfilled places, there is no obvious contractual agreement, and medical schools may wish then to take into account GCSEs, U(K)CAT and BMAT scores, and perhaps other measures such as EPQ and MMI, although the latter two have not yet been validated as predictors of outcome. The use of some sorts of information, such as personal statements and work experience, may however favour more advantaged students [4], although medical schools may wish also to take educational disadvantage into account to widen participation. Ultimately, universities will need to be, “flexible and responsive in their admissions processes” [53].

Whatever medical schools do choose to do (or not to do) the one certainty is that outcomes for the 2020 entry cohort will be followed with interest through their careers, and UKMED will be a major tool in doing that.

### For teachers

Centre assessment grades and rankings may well be problematic for teachers (and recent research suggests that rankings are hard to make for large numbers of candidates [29], and that different teachers use different sorts of information in making their judgements). Since the intention of calculated grades is that they will emulate the actual grades that students would have attained had they taken the exams, then a key emphasis probably has to be on what Petch called “anticipated examination prowess” rather than “scholarly attitude”. How easy it is to do that though is very unclear (and a way to help may be to ask teachers to make both assessments on students, so that differences become clearer). Many teachers will already have provided predicted grades for UCAS applications earlier in the academic year, and whether it is entirely possible to forget those earlier estimates seems unlikely, particularly when much but not all of the information will be the same as earlier. The possibility of parental challenge when centre assessment grades differ from earlier predicted grades may also make the two sets of grades more similar. Cognitive dissonance may additionally make it particularly difficult to forget earlier predictions, although it might be worth remembering the saying often attributed to John Maynard Keynes: “When events change, I change my mind. What do you do?”^n^. Finally, from a research point of view it would be nice to think that information will be collected about how teachers and schools in practice are producing their centre assessment grades, although it seems possible that may not occur due to fears of legal challenge.

### For medical school applicants

Cancellation of A-levels inevitably has produced uncertainty and anxiety for university applicants. Calculated grades have introduced what is to some extent an entirely unknown factor, although on balance it is likely that calculated grades will probably relate closely to predicted grades.

If calculated grades are similar to predicted grades, and given that much selection has in fact already taken place during shortlisting largely, but not entirely, on the basis of predicted grades (and other information such as admissions tests and interviews), then selection this year will mainly be on predicted grades. A particular problem normally is applicants for whom there is under-prediction of grades, as calculated grades are likely also to be under-predicted, but without any actual grades to assess whether that is the case. For those whose grades in normal circumstance are under-predicted, being predicted, say, BBB, but achieving A*AA, there is usually a safety net for a second chance at a place, formally through Clearing or sometimes through other more informal processes such as in medical schools which maintain waiting lists. Quite how, beyond random chance, calculated grades might differ substantially from predicted grades, is unclear. That also means it is difficult to see how Clearing and similar processes will work in 2020. Under-prediction is a particular risk in cases where teachers do not know, or in some cases do not like, their students, perhaps because of attitude, personal characteristics, or other factors, the externality and objectivity of examinations traditionally solving the problem. Petch, once again, put it well, describing,

> “instances, where, in the examination room, candidates have convinced the examiners that they are capable of more than their schools said that they were … Paradoxical as it will seem, examiners are not always on the side of authority; an able rebel can find his wider scope within the so-called cramping confines of an examination.” [3](p.29).

There is a clear echo here of the quote by Yasmin Hussein with which this paper began. Hussein’s concerns are not alone, and the UKMACS study in April 2020 found concerns about fairness were particularly present in medical school applicants from non-selective schools, from Black Asian and Minority Ethnic (BAME) applicants, from female applicants, and from those living in more deprived areas [6].

## Summary

The events of 2020 as a result of the COVID-19 pandemic are extra-ordinary, and unprecedented situations have occurred, of which the cancellations of GCSE and A-level exam cancellations are but one example. The current study should not be seen as criticism of the response of *Ofqual* to that situation; given the circumstances in which it found itself, with examinations cancelled, its solution to the problems has many obvious virtues. For most university applicants there already existed predicted grades from the previous autumn when UCAS applications were submitted, but they would have been on average half a grade or so too high, being aspirational as much as realistic, and also for medical students would have been made by October 2019, whereas calculated grades would be based on teacher predictions in May 2020, albeit with several months of courses missing since March 2020. Raw predicted grades would have wrecked much university planning as numbers of acceptances would inevitably have been far too high. There was also a risk that predicted grades could have been systematically higher from some schools than others – the ones with a tendency to call all of their geese swans -- and that probably applies also to the centre assessment grades to be sent to examination boards. Normalising calculated grades to the broad norms of previous years’ performance, within grades and within schools, circumvents many problems, and the presence of rankings gives the granularity which makes it possible to break the ties which would inevitably arise. Rankings also have the elegant feature that the method ensures that within schools those with higher calculated grades will always have been ranked higher within their school. All of that is admirable and clever, and carefully thought through. It is difficult to see any other solution that could have been adopted given the short time frames and the difficult circumstances. Ofqual probably had little choice either, but to adopt the position that the final grades would be formally equivalent to grades in previous and subsequent years, although it may be problematic.

In official exam statistics the 2020 results will barely be distinguishable from those of other years, the proportions of A*, A, B, etc. by subject and by school not being noticeably different from years before. They will however have been constructed that way, as a sophisticated exercise in what in effect is norm-referencing. Whether calculated grades have the same advantages for candidates and for universities is another matter. This paper has provided evidence that calculated grades will probably not predict future outcomes with the same effectiveness as actual, attained grades, and that is a problem that universities and medical schools and postgraduate deaneries will have to work with, probably for many years as the 2020 cohort works through the system. It seems likely therefore, as Thomson has said, “… this year group will always be different…”[2].

## Data Availability

Researchers wishing to re-analyse the data used for this study can apply for access to the same datasets via UKMED (www.ukmed.ac.uk)

## Acknowledgements

We are grateful to Paul Garrud, Gill Wyness, Paul Newton, Colin Melville and Christian Woodward for their comments on earlier versions of this manuscript, and to Jon Dowell, Peter Tang, Rachel Greatrix and other members of the UKMED Research Group and Advisory Board for their assistance in fast-tracking the paper, and for their comments on it. We also thank Tim Gill for providing us with an unpublished manuscript.

## Contributors

DTS prepared the data extracts, provided details on date sources and variable definitions where required and commented on manuscript drafts. ICM originated the idea for the study, and discussed it with other authors throughout the project. ICM wrote the first draft of the manuscript, and KW, DH, PAT, LP, KYFC and DTS have read, reviewed and commented on earlier drafts and contributed ideas, as well as approving the final draft.

## Funding

KW is a National Institute for Health Research (NIHR) Career Development Fellow (NIHR CDF-2017-10-008) and is principal investigator for the UKMACS and UKMEDP089 projects supported by the NIHR funding.

DH is funded by NIHR grant CDF-2017-10-008 to KW.

PAT’s research time is supported by an NIHR Career Development Fellowship (CDF 2015-08-11), and PAT is also principal investigator for the UKMEDP051 project.

LWP is partly supported by NIHR grant CDF 2015-08-11 to PAT, and a portion of his research time is funded by the UCAT board.

ICM, KYFC and DTS have received no specific funding for this project.

## Disclaimers

KW and DH state that this publication presents independent research funded by the National Institute for Health Research (NIHR). The views expressed are those of the authors and not necessarily those of the NHS, the NIHR or the Department of Health and Social Care.

PAT states this research is supported by an NIHR Career Development Fellowship (CDF 2015-08-11). This paper presents independent research partly funded by the National Institute for Health Research. The views expressed are those of the authors and not necessarily those of the NHS, the NIHR or the Department of Health and Social Care.

KYFC is employed as the Head of Marking and Results at Cambridge Assessment English. The views expressed are those of the authors and do not represent the views of Cambridge Assessment.

DTS is employed by the GMC as a data analyst working on the UKMED project. The views expressed here are his views and not the views of the GMC.

Data sources. UK Medical Education Database (UKMED) UKMEDP051 data extract generated on 13^th^ May 2019. UKMEDP089 extract generated 21^st^ January 2020. UKMEDP112 project, using UKMEDP051 and UKMEDP089 data, approved for publication on 29^th^ May 2020. We are grateful to UKMED for the use of these data. However, UKMED bears no responsibility for their analysis or interpretation.

UKMEDP051 data includes information derived from that collected by the Higher Education Statistics Agency Limited (HESA) and provided to the GMC (HESA Data). Source: HESA Student Record 2002/2003 to 2014/2015. Copyright Higher Education Statistics Agency Limited. The Higher Education Statistics Agency Limited makes no warranty as to the accuracy of the HESA Data, cannot accept responsibility for any inferences or conclusions derived by third parties from data or other information supplied by it.

UKMEDP051 and UKMEDP089 include Universities and Colleges Admissions Service (UCAS) data provided to the GMC (UCAS data). Source: UCAS (application cycles 2007 to 2018). Copyright Universities and Colleges Admissions Service (UCAS). UCAS makes no warranty as to the accuracy of the UCAS Data and cannot accept responsibility for any inferences or conclusions derived by third parties from data or other information supplied by it.

## Competing interests

ICM is a member of the UKMED Research Group and the UKMED Advisory Board, and is also on the UKMACS advisory group.

PAT is a member of the UKMED Research Group. PAT has previously received research funding from the ESRC, the EPSRC, the Department of Health for England, the UCAT Board, and the GMC. In addition, PAT has previously performed consultancy work on behalf of his employing University for the UCAT Board and Work Psychology Group and has received travel and subsistence expenses for attendance at the UCAT Research Group.

KYFC is a member of the UKMED Research Group, and is an employee of Cambridge Assessment - a group of exam boards that owns and administers the BioMedical Admissions Test (BMAT); UK GCSEs and A-levels; and International GCSEs and A-levels.

DTS is a member of the UKMED Research Group and the UKMED Advisory Board and is employed by the GMC as a data analyst working on the UKMED project.

KW, DH and LP declare no competing interests.

## Ethics approval

The authors did not need to seek formal ethical approval for this study as it was a secondary data analysis of existing data. UKMED has received a letter from Queen Marys University of London Ethics of Research Committee on behalf of all UK medical schools to confirm ethics exemption for projects using exclusively UKMED data -https://www.ukmed.ac.uk/documents/UKMED_research_projects_ethics_exemption.pdf

## Provenance and peer review

Not commissioned; reviewed by the UKMED Research Subgroup and Advisory board, and to submitted to an externally peer reviewed journal. This paper was fast-tracked through the UKMED governance processes as with other COVID-19 related research projects elsewhere.

## Footnotes

a SQA Highers are taken the year before (rather like AS-levels used to be) and therefore they will be available for 2020 applicants. Advanced Highers will not be available and will be estimated.

b A number of students, designated as ‘external’ or ‘private’, do not have regular teachers who can make such assessments and Ofqual has announced that they will not be able to receive calculated grades. Cases may include students taking a gap year and retaking A-levels, or a student studying GCSE or A-level German at a *Samstagsschule* who for administrative convenience takes the exam at a local school where the subject is not actually taught. There no doubt are many other A-level candidates affected as well.

c A parallel may be seen in the phrase still seen on UK banknotes that “I promise to pay the bearer on demand the sum of ten pounds”, originally referring to a particular amount of gold. A bank note and gold may then have been equivalent in legal status for paying bills, but clearly were not equivalent in other properties, such as melting point, flammability, etc..

d https://www.bbc.co.uk/news/education-44525719

e https://www.hepi.ac.uk/2019/08/14/pqa-just-what-does-it-mean/

f In some studies a scoring of A*=6, A=5, B=4 is used. The 12,10,8… scoring was introduced so that AS levels, weighted at half an A-level, could be scored as A=5, B=4 etc (there being no A* grade at AS-level). For most purposes A*=12, A=10 … is equivalent in all respects to A*=6, A=5, etc, apart from a scaling factor.

g https://www.ucas.com/advisers/managing-applications/predicted-grades-what-you-need-know [Accessed 13th April 2020].

h https://ukmacs.wordpress.com/

i An upload for P51 dated 20th April 2020 is now available but pre-processing is required before the data can be analysed properly.

j See https://www.gov.uk/government/publications/reliability-of-assessment-compendium for a range of important papers commissioned and published by Ofqual.

k https://foundationprogramme.nhs.uk/wp-content/uploads/sites/2/2019/11/UKFP-2020-EPM-Framework-Final-1.pdf,

l In part this reflects the fact that some students, particularly weak ones, are given an EPM score, but then fail finals.

m This is based on deciles being calculated in a way that are equated to levels used in the present calculation. Of course if calculated strictly as deciles then of necessity 10% will still remain in the top decile, etc.. That difficulty of deciles will in large part be removed when the 2020 entrants graduate as UKMLA should be on stream by then.

n The phrase was probably said in 1970 by Paul Samuelson, a Nobel-prize winning economist, although in 1978 he himself attributed it to Keynes. https://quoteinvestigator.com/2011/07/22/keynes-change-mind/

## Notes

### Author Declarations

The authors did not need to seek formal ethical approval for this study as it was a secondary data analysis of existing data. UKMED has received a letter from Queen Marys University of London Ethics of Research Committee on behalf of all UK medical schools to confirm ethics exemption for projects using exclusively UKMED data - https://www.ukmed.ac.uk/documents/UKMED_research_projects_ethics_exemption.pdf

